# Clinical and Economic Impact of COVID-19 on Plantation Workers: Preliminary Results from the Guatemala Agricultural Workers and Respiratory Illness Impact (AGRI) Study

**DOI:** 10.1101/2022.02.07.22270274

**Authors:** Daniel Olson, Diva M. Calvimontes, Molly M. Lamb, Gerber Guzman, Edgar Barrios, Andrea Chacon, Neudy Rojop, Kareen Arias, Melissa Gomez, Guillermo Antonio Bolanos, Jose Monzon, Anna N. Chard, Chelsea Iwamoto, Lindsey M. Duca, Nga Vuong, Melissa Fineman, Kelsey Lesteberg, David Beckham, Mario L. Santiago, Kendra Quicke, Gregory Ebel, Emily Zielinski Gutierrez, Eduardo Azziz-Baumgartner, Frederick G. Hayden, Hani Mansour, Kathryn Edwards, Lee S. Newman, Edwin J. Asturias

## Abstract

We evaluated the clinical and socioeconomic burdens of respiratory disease in a cohort of Guatemalan banana plantation workers. All eligible workers were offered enrollment from June 15–December 30, 2020, and annually, then followed for influenza-like illnesses (ILI) through: 1) self-reporting to study nurses, 2) sentinel surveillance at health posts, and 3) absenteeism. Workers with ILI submitted nasopharyngeal swabs for influenza, RSV, and SARS-CoV-2 testing, then completed surveys at days 0, 7, and 28. Through October 10, 2021, 1,833 workers developed 169 ILIs (12.0/100 person-years) and 43 (25.4%) of these ILIs were laboratory-confirmed SARS-CoV-2 (3.1/100 person-years). Workers with SARS-CoV-2-positive ILI reported more anosmia (p<0.01), dysgeusia (p<0.01), difficulty concentrating (p=0.01), and irritability (p=0.01), and greater clinical and well-being severity scores (Flu-iiQ) than test-negative ILIs; they also had greater absenteeism (p<0.01) and lost income (median US$127.1, p<0.01). These results support the prioritization of Guatemalan farm workers for COVID-19 vaccination.

## INTRODUCTION

Essential workers have been at greater risk of COVID-19 compared to the general population, but little is known about the risk to people working within the agricultural sector in low-middle-income countries (LMICs) (1-3). Though limited data from the United States (U.S.) have demonstrated a high SARS-CoV-2 burden in this population (1), many agricultural workers continued working throughout the pandemic (4). In LMICs, agricultural workers play a critical role in food security and represent a significant economic force. In Guatemala, they comprise 35% of the overall labor force, and agricultural products account for 45% of all exports and 11.3% of total gross domestic product (5). Guatemala is also a major trading partner of the U.S., exporting US$ 2.1 billion in agricultural products annually, including nearly 50% of the U.S. banana supply (6). Therefore, agricultural workers in Guatemala and similar LMICs are arguably “essential” not only for local food security, but also the food security of international trading partners such as the U.S.

In addition to increased risk of exposure to SARS-CoV-2, the virus that causes COVID-19, people working in the agricultural sector may also be at increased risk of poor clinical outcomes from COVID-19 because of a high prevalence of comorbidities associated with environmental stress, such as chronic kidney disease of unknown origin (CKDu, “Mesoamerican nephropathy”) (7-9). Economic outcomes, such as work absenteeism and decreased job performance while working (“presenteeism”), are also likely significant, as is the case with influenza (10-14). As agricultural workers are often the primary income earners for their households, the consequences may extend to their households and communities. Despite the increased clinical and economic vulnerability of agricultural workers, and their critical role in global food security, little is known about the socioeconomic consequences of COVID-19 and other respiratory illnesses among this essential workforce, and the subsequent impacts on their households and communities.

The “<u>AG</u>ricultural workers and <u>R</u>espiratory Illness <u>I</u>mpact (AGRI) Study” was initially designed as an influenza cohort and expanded to include other viral respiratory pathogens, including SARS-CoV-2. The study has two primary aims: 1) to characterize the clinical and the socioeconomic outcomes of acute respiratory viral infections among Guatemalan plantation workers, and 2) to measure the effectiveness of a workplace-based vaccination program in improving these outcomes. This paper provides a comprehensive description of the AGRI cohort and a summary of clinical and economic outcomes from our first year of virologic surveillance.

## METHODS

### Study Setting and Population

The 5-year study is being conducted within a large banana plantation in the coastal lowlands of southwest Guatemala. The farm workers are exposed to high temperatures and humidity, and are at risk for environment-associated chronic medical conditions such as CKDu (7, 15) . Previous surveys (2015, 2017-18) found a predominantly young, male, and economically vulnerable workforce, in which the farmworkers are typically the sole income earners for their households and report high rates of food insecurity, similar to other agribusiness workers in the region and migrant worker populations in the U.S. (16, 17). The regional population experiences high levels of food insecurity, stunting, poverty, and communicable diseases, and low access to healthcare (18, 19).

As is typical in many agribusinesses, field workers and packaging workers are given a baseline pay, with daily bonuses based on their productivity recorded by the company. Managers and workers in administrative job categories are paid by day. If a worker develops illness and is provided an excused absence by their manager, they receive 2/3 of their baseline pay for the duration of their excused absence, up to a maximum of US$ 15.6 per day. Workers with laboratory-confirmed SARS-CoV-2 are mandated to quarantine at home with excused absences for up to two weeks duration.

All eligible workers within the nine banana plantation worksites were offered enrollment in the study from June 15^th^ to December 30^th^, 2020, and annually. Inclusion criteria include age ≥18 years, plans to remain employed by the agribusiness ≥1 year, access to a telephone, and agreement to allow use of company-based absenteeism and job performance records. For this analysis, participant follow-up was performed through October 10, 2021, and all study procedures (testing, follow-up) performed after this date were considered missing, even if the associated ILI case was identified prior.

Following written informed consent, study nurses collected contact information as well as demographic, occupational, socioeconomic, and clinical data, including risk factors for severe COVID-19. Workers provided enrollment and annual blood specimens that will be screened for markers of chronic kidney disease (estimated glomerular filtration rate, (20)), anti-SARS-CoV-2 nucleocapsid IgG (Roche Elecsys^®^ immunoassay), and in some cases, anti-SARS-CoV-2 neutralizing antibodies (Beckham/Santiago Laboratories, University of Colorado, Aurora, CO, USA). Workers leaving employment have exit interviews and are removed from the study, but data collected during their employment are retained in the study database.

### Surveillance for Influenza-like Illness (ILI)

Following enrollment, all workers began prospective active surveillance for influenza-like illness (ILI), initially defined as self-reported fever/temperature ≥38°C *and* cough in the last 10 days to focus on detection of influenza (21). In January 2021, the ILI case definition was expanded to include presence of fever, cough, *or* shortness of breath in the last 10 days (COVID-19-like illness (22)), to increase sensitivity of COVID-19 case detection (23).

The study employed three strategies for detecting ILI: 1) symptom screening through workers self-reporting symptoms to a study nurse during weekly worksite visits, work supervisors screening workers daily for cough and fever, and telephone contact to a study nurse by workers experiencing symptoms at any time; 2) sentinel surveillance of all workers presenting to worker health posts within the plantation with ILI; and 3) active monitoring and ILI screening phone calls to absent workers identified on the company absenteeism registry. In February 2021, absenteeism calls were discontinued because these cases of ILI were consistently identified through other surveillance approaches.

### Syndromic Illness Characterization

Workers with ILI were interviewed by study nurses and provided clinical, epidemiologic, and outcome data for themselves and general epidemiologic and socioeconomic outcome data for their households. Study nurses also collected a nasopharyngeal (NP) swab, which was placed in viral transport media and tested within 24 hours of specimen collection for SARS-CoV-2 using the Q COVID rapid antigen test (Q-NCOV-01G, SD Biosensor^®^, Republic of Korea) (24). Aliquots were also tested for influenza A/B and respiratory syncytial virus (RSV) using the Roche cobas^®^ Liat Influenza A/B (& RSV) real-time polymerase chain reaction (rtRT-PCR) instrument (25). ILI cases who tested positive for SARS-CoV-2, influenza, or RSV are hereafter referred to as SARS-CoV-2-positive ILI, influenza-positive ILI, and RSV-positive ILI, respectively. A subset of available ILI specimens collected through April 2021 (n=40) were also tested for an additional 15 respiratory pathogens using the multiplex BioFire FilmArray RP2.1 assay (26).

Viral testing results were shared with participants when available (usually within 24 hours) and weekly with the Guatemala Ministry of Health.

### Clinical and Socioeconomic Outcome Assessments

The study relied on a case-cohort study design to measure self-reported clinical and socioeconomic outcomes. All subjects in the overall cohort with ILI were considered “cases.” Each week, a sub-cohort of 15 enrolled workers without ILI in the preceding 28 days were selected at random (∼5% of the cohort per month) and considered “controls.” Follow-up surveys were administered by study nurses over the phone to cases at 1 and 4 weeks following their ILI visit. Controls were notified that they had been selected (day 0) and were then provided the same surveys 1 and 4 weeks later; controls did not undergo diagnostic testing, and a control who developed ILI during the 4-week follow-up was considered a case at the time of illness.

Clinical and well-being outcomes were collected using the Flu-iiQ™ inventory (27), which is a validated Spanish-language outcome measure designed for clinical and epidemiologic outpatient studies of influenza and RSV. The inventory includes 13 items for symptom severity, which comprise a combined “systemic score” (7 items) and “respiratory score” (6 items). The well-being scores include an “impact on daily activities score” (7 items), “impact on emotions score” (4 items), and “impact on others score” (5 items). Each combined score is averaged by the number of individual items such that all scores are between 0 and 3, with a higher score indicating greater severity or negative impact on well-being. The follow-up surveys also collected health-seeking behavior (e.g., hospitalization, medication usage, etc.).

During the follow-up surveys, economic outcomes were assessed using questions adapted from the 2016 WHO Manual for Estimating the Economic Burden of Seasonal Influenza (28) and supplemented with the World Bank National Survey of Living Conditions (29), which includes a Spanish translation (ENCOVI) previously used in Guatemala (30), and compared to the basic food basket price in Guatemala, which reflects the minimum kilocalorie (2,262 kilocalorie) intake for a 4.77-member household for one month (US$ 386.3 in March 2021 (31)).The survey collected data on direct medical costs, direct non-medical costs (i.e., transportation), and indirect costs related to loss of productivity (i.e., absenteeism) for both the worker and household. Though not included in this analysis, company reported individual-level data will be linked to workers, including absenteeism, productivity metrics (task-specific units of production such as tons of bananas harvested/day), and wages.

### Statistical Analysis

Incidence density (number of cases per person-time of follow-up) of ILI and pathogen-specific ILI were calculated. Descriptive statistics were used to calculate the differences between clinical and socioeconomic outcomes between groups. For normally distributed continuous variables, means and standard deviations were calculated, and Student’s T-tests were used to determine significant differences between groups. For non-normally distributed continuous variables, medians and interquartile ranges were calculated, and the Wilcoxon Rank-Sum test was used to determine significant differences between groups. For categorical variables, chi-square and Fischer’s exact tests were used to determine significant differences in distribution of categories between groups. For all analyses, a p-value <0.05 was considered statistically significant.

### Ethical Oversight

The study was approved by the Colorado Multiple Institutional Review Board (COMIRB protocol #19-1836) and the Guatemala Ministry of Health National Ethics Committee (HRMC-560-2020). The local Southwest Trifinio Community Advisory Board for Research agreed to the study. Workers receive no compensation for study participation.

## RESULTS

Between June 15, 2020 and October 10, 2021, 2,371 workers were screened for enrollment; 160 (6.7%) were ineligible and 378 (17.1%) declined participation (**Figure 1**). Of the 1,833 enrolled participants, 1,590 (86.7%) remained active in the study as of October 10, 2021, representing 1,402.9 person-years of surveillance. Workers who declined participation were slightly younger (29.6 vs 30.9 years, p<0.01) than participants but had similar sex distribution and ethnicity.

**Figure 1.**
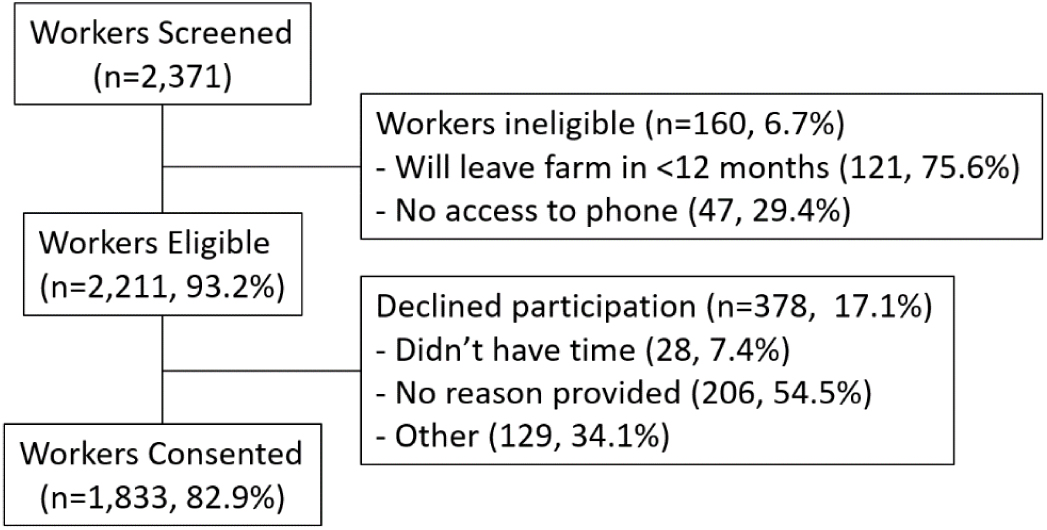
CONSORT Diagram. Summary of cohort enrolled in the AGRI Study in SW Guatemala from June 2020 to October 2021, and followed up through October 10, 2021. Ineligible and non-consenting workers were able to provide multiple reasons for not participating. Only workers who completed the Day 0 (diagnosis) visit were called on Day 7 and Day 28. Follow up visits scheduled for after October 10th, 2021 were considered missing.

The enrolled cohort characteristics are summarized in **Table 1**. Most workers were male (84.1%) and worked in the fields (69.0%). Self-reported chronic medical conditions were uncommon except for obesity (body mass index [BMI] ≥30 kg/m^2^, 11.3%) and kidney disease(3.2%); 12.8% of workers (n=234) took medications, the majority of whom (n=122, 52%) took vitamins, followed by pain relievers/anti-inflammatories (14%), antibiotics (7%), diabetes-related medication (7%), and proton pump inhibitors (6%). Only 5.9% reported ever having received an influenza vaccination, including 17 (6.4%) of the 267 workers who self-reported chronic diseases. Workers began to receive COVID-19 vaccination through the workplace in August 2021 (ChAdOx1, AstraZeneca, England; and mRNA-1273, Moderna, USA). Of 1,334 workers enrolled in 2020 with samples available, 616 (46.2%) were SARS-CoV-2 anti-nucleocapsid IgG reactive.

**Table 1.** Characteristics of cohort participants, Guatemala (n=1,833)

|  |  |
| --- | --- |
| **Worker Demographics** |  |
| Age in years (mean, SD) | 30.9 (8.7) |
| Male sex, n (%) | 1541 (84.1) |
| Ethnicity, n (%): Ladino | 801 (43.7) |
| Indigenous | 113 (6.2) |
| Other | 3 (0.2) |
| Don't know | 916 (43.7) |
| **Worker Health, n (%)** |  |
| Obesity (measured BMI > 30 kg/m<sup>2</sup>), n= 1159 with data | 131 (11.3) |
| Class 1 (BMI 30-<35 kg/m<sup>2</sup>) | 103 |
| Class 2 (BMI 35-<40 kg/m<sup>2</sup>) | 24 |
| Class 3 (BMI ≥40 kg/m<sup>2</sup>) | 4 |
| Kidney disease | 58 (3.2) |
| Blood disorder (e.g., sickle cell disease) | 25 (1.4) |
| Cardiovascular disease (e.g., heart failure, CAD) | 29 (1.6) |
| Diabetes | 27 (1.5) |
| Liver disease | 19 (1.0) |
| Asthma | 10 (0.6) |
| Pulmonary Disease (e.g., COPD) | 10 (0.6) |
| Neurologic disease (e.g., stroke) | 10 (0.6) |
| Taking medications | 234 (12.8) |
| Received influenza vaccine | 108 (5.9) |
| **Work Conditions** |  |
| Type of Work, n (%) |  |
| Administration | 60 (3.3) |
| Field worker | 1264 (69.0) |
| Field manager | 77 (4.2) |
| Packer (plant worker) | 413 (22.5) |
| Plant manager | 19 (1.0) |
| Duration of employment, n (%) |  |
| ≤2 years | 1115 (60.9) |
| 3-4 years | 242 (13.2) |
| ≥5 years | 475 (25.9) |
| Monthly income, \$USD, median (IQR) | 337.2 (311.3, 389.1) |
| **Household Conditions** |  |
| # adults in house, median (IQR) | 3 (2-4) |
| # children in house, median (IQR) | 2 (1-3) |
| Concern about food insecurity in last year, n (%) | 1063 (58.0) |
| Household monthly income, \$USD, median (IQR) | 363.2 (324.3-505.8) |
| \$USD spent in the last 7 days, median (IQR): | |
| Meat, fish, and seafood | 25.9 (13.0-38.9) |
| Milk, eggs, and dairy products | 15.6 (9.7-25.9) |
| Greens, vegetables, and fruit | 13.0 (6.8-19.5) |
| Alcoholic drinks and tobacco | 0 (0-0) |

Abbreviations: SD=standard deviation, BMI=body mass index, CAD=coronary artery disease, IQR=interquartile range, USD=US dollars

Household size averaged 5.7 individuals (3.3 adults, 2.3 children) with nearly half the workers (n=877; 48.2%) living in the urban municipality of Coatepeque; the study catchment area is approximately 2,600 km^2^ (**Figure 2**). Median self-reported monthly income for the individual worker was US$ 337.2 (interquartile range [IQR]=311.3, 389.1) and for the household was US$ 363.2 (IQR=324.3, 505.8); 58.0% of workers reported being worried about the inability to purchase food in the preceding 12 months.

**Figure 2.**
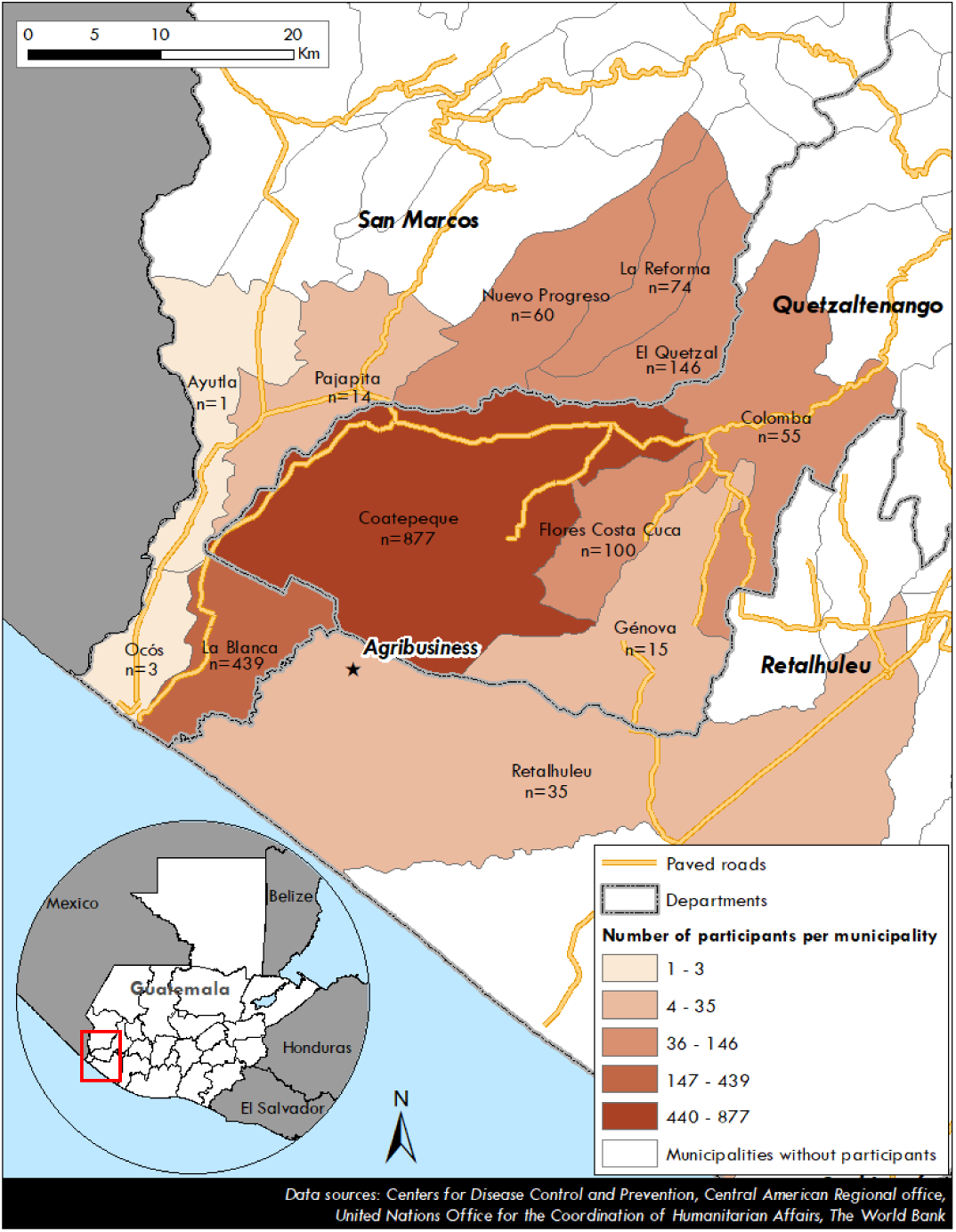
Map of the study region (2,600 km^2^) showing number of enrolled subjects living in each municipality, n=1,819 with reported data.

### Asymptomatic controls

Of the 915 asymptomatic controls that were randomly selected (August 10, 2020– October 10, 2021), the study team was able to contact 696 (76.0%) by phone. There were no significant differences in enrollment characteristics between those contacted and not contacted. Of the 696 controls who were contacted initially, 623 (89.5%) and 588 (84.4%) were successfully contacted at 1 and 4 weeks, respectively.

### Absenteeism

From August 31, 2020 to February 19, 2021, 736 workers (51.4%) had ≥1 day of work absence. Study personnel contacted 504 (68.5%) after three attempts, and there were no differences between contacted and uncontacted workers other than number of children (2.7 vs 2.2, respectively, p<0.01). Risk associations for absenteeism are shown in **Supplemental Table 1**.

### Respiratory Illnesses

Between June 15, 2020 and October 10, 2021, the study identified 169 ILI episodes occurring among 145 unique persons; of those, 136 (93.8%) persons (for 157 ILI episodes) and 129 (89.0%) persons (for 149 ILI episodes) completed the 7- and 28-day follow up surveys by analysis closeout (**Supplemental Table 2**). Of the 153 ILI episodes (among 132 unique persons) with completed SARS-CoV-2 antigen testing by analysis closeout, 43 (28.1%) were positive for SARS-CoV-2. Of 151 ILI episodes (among 131 unique persons) with complete influenza and RSV RT-PCR testing, 6 (3.7%) were RSV-positive, and 0 were influenza-positive. The incidence density of ILI was 12.0 person-years (P-Y) and of SARS-CoV-2 was 3.1/100 P-Y (**Figure 3**).

**Table 2.** Comparison of Self-Reported Outcomes in SARS-CoV-2-positive ILI vs SARS-CoV-2-negative ILI.

|  | Day 0<br>SARS-CoV-2<br>positive<br>(n=43) | Day 0<br>SARS-CoV-2<br>negative<br>(n=110) | p-value* | Day 7<br>SARS-CoV-2<br>positive<br>(n=41) | Day 7<br>SARS-CoV-2<br>negative<br>(n=103) | p-value* | Day 28<br>SARS-CoV-2<br>positive<br>(n=38) | Day 28<br>SARS-CoV-2<br>negative<br>(n=97) | p-value* |
| --- | --- | --- | --- | --- | --- | --- | --- | --- | --- |
| **Clinical Symptoms** |  |  |  |  |  |  |  |  |  |
| # days of fever (if fever), mean (SD) | 3.3 (2.0) | 2.3 (1.5) | <0.01 | 4.4 (2.8) | 3.3 (2.5) | 0.08 | 0.5 (0.0) | 3.4(2.2) | 0.04 |
| # days of cough (if cough), mean (SD) | 3.3 (1.9) | 3.6 (2.1) | 0.45 | 6.1 (2.3) | 5.1 (2.6) | 0.07 | 4.5 (4.9) | 5.1 (3.5) | 0.84 |
| Have you felt the following (last 24 hours), n (%): |  |  |  |  |  |  |  |  |  |
| Fever | 32 (74.4) | 60 (54.6) | 0.02 | 2 (4.9) | 11 (10.8) | 0.35 | 4 (10.5) | 4 (4.2) | 0.22 |
| Nasal Congestion | 18 (41.9) | 39 (35.5) | 0.46 | 5 (12.2) | 17 (16.7) | 0.50 | 2 (5.3) | 6 (6.3) | 1.00 |
| Myalgia | 23 (53.5) | 46 (41.8) | 0.19 | 3 (7.3) | 10 (9.8) | 0.76 | 5 (13.2) | 6 (6.3) | 0.19 |
| Headache | 30 (69.8) | 55 (50.0) | 0.03 | 13 (31.7) | 21 (20.6) | 0.16 | 8 (21.1) | 14 (14.6) | 0.36 |
| Cough | 24 (55.8) | 39 (35.5) | 0.02 | 7 (17.5) | 14 (13.7) | 0.57 | 3 (7.9) | 7 (7.3) | 1.00 |
| Sore Throat | 15 (34.9) | 50 (45.5) | 0.23 | 6 (14.6) | 14 (13.7) | 0.89 | 4 (10.5) | 7 (7.3) | 0.51 |
| Dysgeusia | 21 (48.8) | 27 (24.6) | <0.01 | 8 (19.5) | 6 (5.9) | 0.01 | 3 (7.9) | 2 (2.1) | 0.14 |
| Expectoration | 16 (37.2) | 45 (40.9) | 0.67 | 12 (29.3) | 18 (17.7) | 0.12 | 7 (10.5) | 8 (8.3) | 0.74 |
| Fatigue | 24 (55.8) | 30 (27.3) | <0.01 | 7 (17.1) | 11 (10.8) | 0.31 | 6 (15.8) | 5 (5.2) | 0.04 |
| Anosmia | 19 (44.2) | 19 (17.3) | <0.01 | 9 (22.0) | 3 (2.9) | <0.01 | 3 (7.9) | 2 (2.1) | 0.14 |
| Loss of Appetite | 17 (39.5) | 31 (28.2) | 0.17 | 4 (9.8) | 5 (4.9) | 0.28 | 1 (2.6) | 2 (2.1) | 1.00 |
| Dyspnea | 13 (30.2) | 25 (22.9) | 0.35 | 4 (9.8) | 5 (4.9) | 0.28 | 4 (10.8) | 1 (1.0) | 0.02 |
| Neck Pain | 14 (33.3) | 18 (16.4) | 0.02 | 6 (14.6) | 5 (4.9) | 0.05 | 4 (10.5) | 4 (4.2) | 0.22 |
| Interrupted Sleep | 11 (25.6) | 19 (17.3) | 0.24 | 8 (19.5) | 6 (5.9) | 0.01 | 3 (7.9) | 4 (4.2) | 0.40 |
| Wheezing | 4 (9.5) | 14 (12.7) | 0.78 | 1 (2.4) | 5 (4.9) | 0.67 | 4 (10.5) | 1 (1.0) | 0.02 |
| **Well-Being\*\*** |  |  |  |  |  |  |  |  |  |
| Have you had difficulty with (last 24 hrs), n (%): |  |  |  |  |  |  |  |  |  |
| Getting out of bed | 16 (37.2) | 26 (23.6) | 0.09 | 5 (12.2) | 4 (3.9) | 0.12 | 2 (5.3) | 4 (4.2) | 1.00 |
| Preparing meals... | 8 (18.6) | 10 (9.1) | 0.10 | 3 (7.3) | 1 (1.0) | 0.07 | 2 (5.3) | 2 (2.1) | 0.32 |
| Performing usual... | 16 (37.2) | 27 (24.6) | 0.12 | 4 (9.8) | 3 (2.9) | 0.10 | 3 (7.9) | 3 (3.1) | 0.35 |
| Leaving the home... | 13 (30.2) | 14 (12.7) | 0.01 | 6 (14.6) | 0 (0) | <0.01 | 2 (5.3) | 2 (2.1) | 0.32 |
| Concentrating on... | 18 (41.9) | 23 (20.9) | 0.01 | 4 (9.8) | 2 (2.0) | 0.06 | 2 (5.3) | 5 (5.2) | 1.00 |
| Taking care of... | 16 (37.2) | 13 (11.7) | <0.01 | 4 (9.8) | 0 (0) | 0.01 | 2 (5.3) | 4 (4.2) | 1.00 |
| Leave the room | 9 (20.9) | 14 (12.7) | 0.20 | 4 (9.7) | 0 (0) | 0.01 | 2 (5.3) | 2 (2.1) | 0.32 |
| Have you felt the following (last 24 hours), n (%): |  |  |  |  |  |  |  |  |  |
| Irritable | 25 (58.1) | 37 (33.6) | 0.01 | 4 (9.8) | 8 (7.8) | 0.74 | 4 (10.5) | 6 (6.3) | 0.47 |
| Defenseless | 18 (41.9) | 20 (18.2) | <0.01 | 5 (12.2) | 5 (4.9) | 0.12 | 3 (7.9) | 3 (3.1) | 0.35 |
| Worried | 19 (44.2) | 35 (31.8) | 0.15 | 13 (31.7) | 8 (7.8) | <0.01 | 5 (13.2) | 10 (10.4) | 0.65 |
| Frustrated | 15 (34.9) | 16 (14.6) | <0.01 | 3 (7.3) | 6 (5.9) | 0.72 | 2 (5.3) | 5 (5.2) | 1.00 |
| People worrying... | 30 (69.8) | 57 (51.8) | 0.04 | 18 (43.9) | 21 (20.6) | <0.01 | 11 (29.0) | 18 (18.8) | 0.20 |
| Being a burden | 13 (30.2) | 30 (27.3) | 0.71 | 9 (22.0) | 10 (9.8) | 0.05 | 5 (13.2) | 9 (9.4) | 0.52 |
| People being... | 13 (30.2) | 24 (21.8) | 0.27 | 8 (19.5) | 8 (7.8) | 0.05 | 7 (18.4) | 7 (8.3) | 0.10 |
| Needing to depend... | 16 (37.2) | 23 (20.9) | 0.04 | 10 (24.4) | 9 (8.8) | 0.01 | 6 (15.8) | 10 (10.4) | 0.39 |
| People having to do extra... | 15 (34.9) | 21 (19.1) | 0.04 | 10 (24.4) | 10 (9.8) | 0.02 | 8 (21.1) | 8 (8.3) | 0.04 |
| **Flu-iiQ Severity Scores\*\*\*** |  |  |  |  |  |  |  |  |  |
| Systemic Score | 0.87 (0.68) | 0.52 (0.55) | <0.01 | 0.21 (0.35) | 0.12 (0.24) | 0.19 | 0.13 (0.31) | 0.09 (0.28) | 0.41 |
| Respiratory Score | 0.50 (0.43) | 0.47 (0.45) | 0.64 | 0.16 (0.28) | 0.16 (0.32) | 0.99 | 0.14 (0.4) | 0.08 (0.21) | 0.41 |
| Impact on Daily Activities | 0.50 (0.85) | 0.22 (0.40) | 0.01 | 0.14 (0.40) | 0.02 (0.08) | 0.05 | 0.06 (0.21) | 0.04 (0.20) | 0.71 |
| Impact on Emotions Score | 0.62 (0.57) | 0.31 (0.45) | <0.01 | 0.17 (0.34) | 0.07 (0.22) | 0.09 | 0.20 (0.24) | 0.08 (0.23) | 0.60 |
| Impact on Others Score | 0.65 (0.65) | 0.44 (0.64) | 0.08 | 0.37 (0.52) | 0.16 (0.40) | 0.03 | 0.25 (0.47) | 0.16 (0.41) | 0.27 |
| **Epidemiology** |  |  |  |  |  |  |  |  |  |
| # in house w/ similar illness (last 2 weeks) | 0.30 (0.64) | 0.13 (0.36) | 0.10 | 0.34 (0.62) | 0.17 (0.49) | 0.09 | 0.11 (0.39) | 0.14 (0.41) | 0.61 |
| Index case in house, n (%) |  |  |  |  |  |  |  |  |  |
| Self | 35 (81.4) | 97 (88.2) |  | 34 (82.9) | 96 (94.1) |  | 33 (86.8) | 90 (93.8) |  |
| Spouse | 2 (4.7) | 3 (2.7) |  | 1 (2.4) | 1 (1.0) |  | 1 (2.6) | 1 (1.0) |  |
| Parent | 0 (0) | 2 (1.8) | 0.13 | 1 (2.4) | 1 (1.0) | 0.10 | 0 (0) | 0 (0) | 0.24 |
| Sibling | 3 (7.0) | 3 (2.7) |  | 3 (7.3) | 2 (2.0) |  | 3 (7.9) | 3 (3.1) |  |
| Cousin | 1 (2.3) | 0 (0) |  | 1 (2.4) | 0 (0) |  | 0 (0) | 0 (0) |  |
| Child | 0 (0) | 4 (3.6) |  | 0 (0) | 1 (1.0) |  | 0 (0) | 2 (2.1) |  |
| *p-values are from t-tests for continuous variables, and chi-square test for categorical variables.<br>**Items truncated per Flu-iiQ licensing agreement; full items available from the author.<br>***Higher scores indicate worse outcomes |  |  |  |  |  |  |  |  |  |

**Figure 3.**
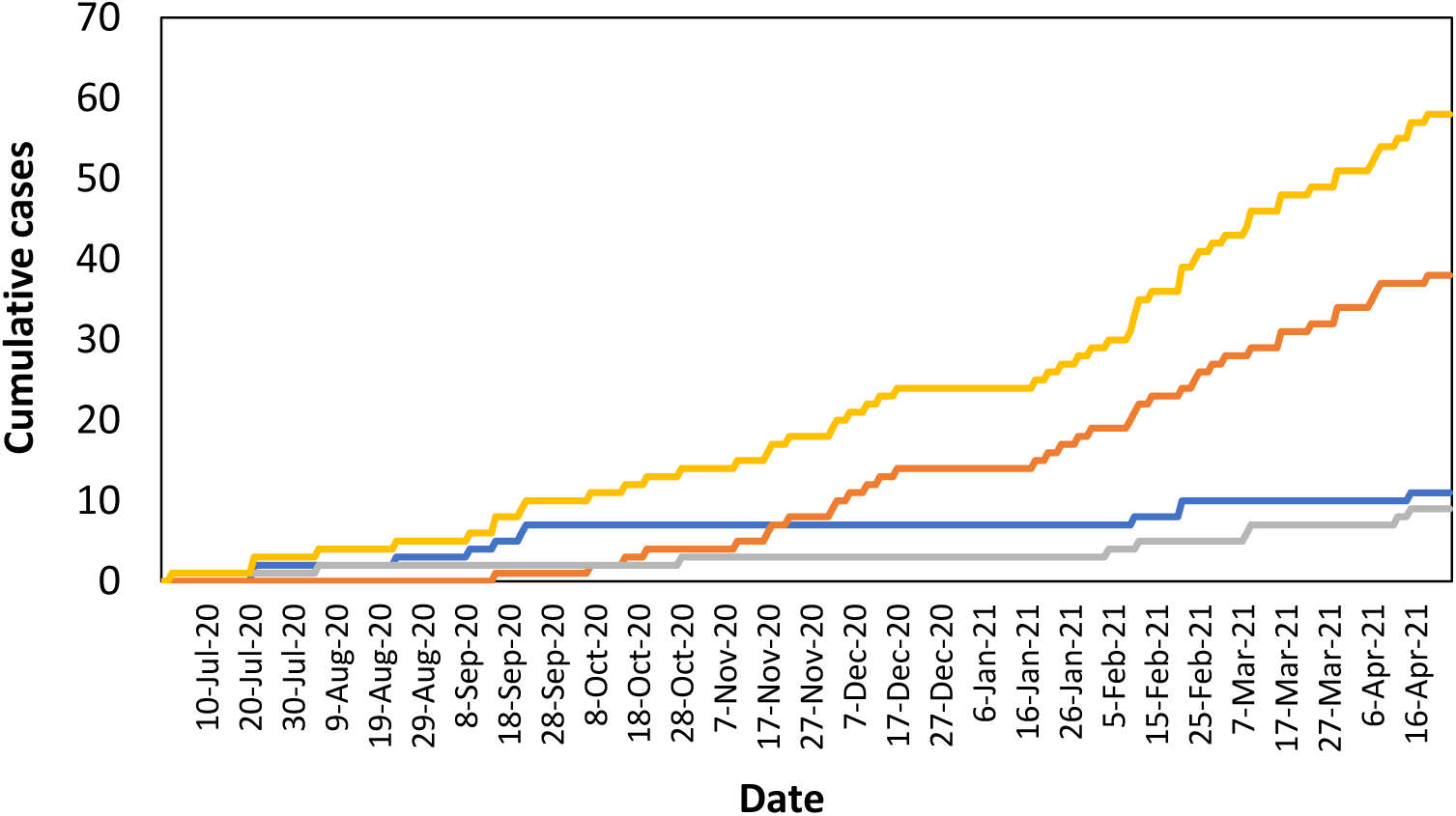
Cumulative ILI Events in the AGRI Cohort, June 15, 2020 to October 10, 2021. From June 2020–October 2021, ILI was defined as “cough and fever”; in January 2021, the ILI case definition was expanded to “cough or fever or shortness of breath”. Includes all-cause ILI (yellow), SARS-CoV-2-positive ILI (blue), SARS-CoV-2 negative ILI (orange), and ILI without test obtained (gray).

BioFire FilmArray RP2.1 testing (n=40) on available specimens confirmed 9 of 9 SARS-CoV-2 infections tested, 1 of 1 RSV infection, and identified an additional 8 picornaviruses (rhino/enterovirus target on FilmArray) and 6 seasonal coronavirus (3 NL63, 1 OC43, and 2 N229E) ILI cases. The adult worker was usually the index case within the household for both SARS-CoV-2 ILI (>80%) infections (**Table 2**) and ILI (>85%) (**Supplemental Table 2**).

Workers with SARS-CoV-2 had longer fever duration at the time of diagnosis (day 0; 3.3 vs 2.3 days; p<0.01) and increased frequency of anosmia (44.2% vs 17.3%; p<0.01) and dysgeusia (48.8% vs 24.6%; p<0.01), compared to SARS-CoV-2-negative workers with ILI (**Table 2**). SARS-CoV-2 cases were also more likely to have difficulty concentrating (41.9% vs 20.9%, p=0.01), irritability (58.1% vs 33.6%, p=0.01), and dependence on others (37.2% vs 20.9%, p=0.04). SARS-CoV-2-positive workers had higher systemic Flu-iiQ severity scores (indicating greater disease severity) at diagnosis than SARS-CoV-2-negative workers, but other clinical scores remained non-significant. SARS-CoV-2-positive workers reported worse impact on “daily activities” (0.50 vs 0.22, p=0.01) and “emotions” (0.62 vs 0.31, p<0.01) scores than SARS-CoV-2-negative workers at diagnosis, and worse “impact on others” score at day 7 (0.37 vs 0.16, p=0.03), with all other Flu-iiQ well-being scores showing a similar non-significant trend (**Table 2, Figure 4**). Clinical outcomes of workers with ILI episodes versus asymptomatic controls are shown in **Supplementary Table 2**.

**Figure 4.**
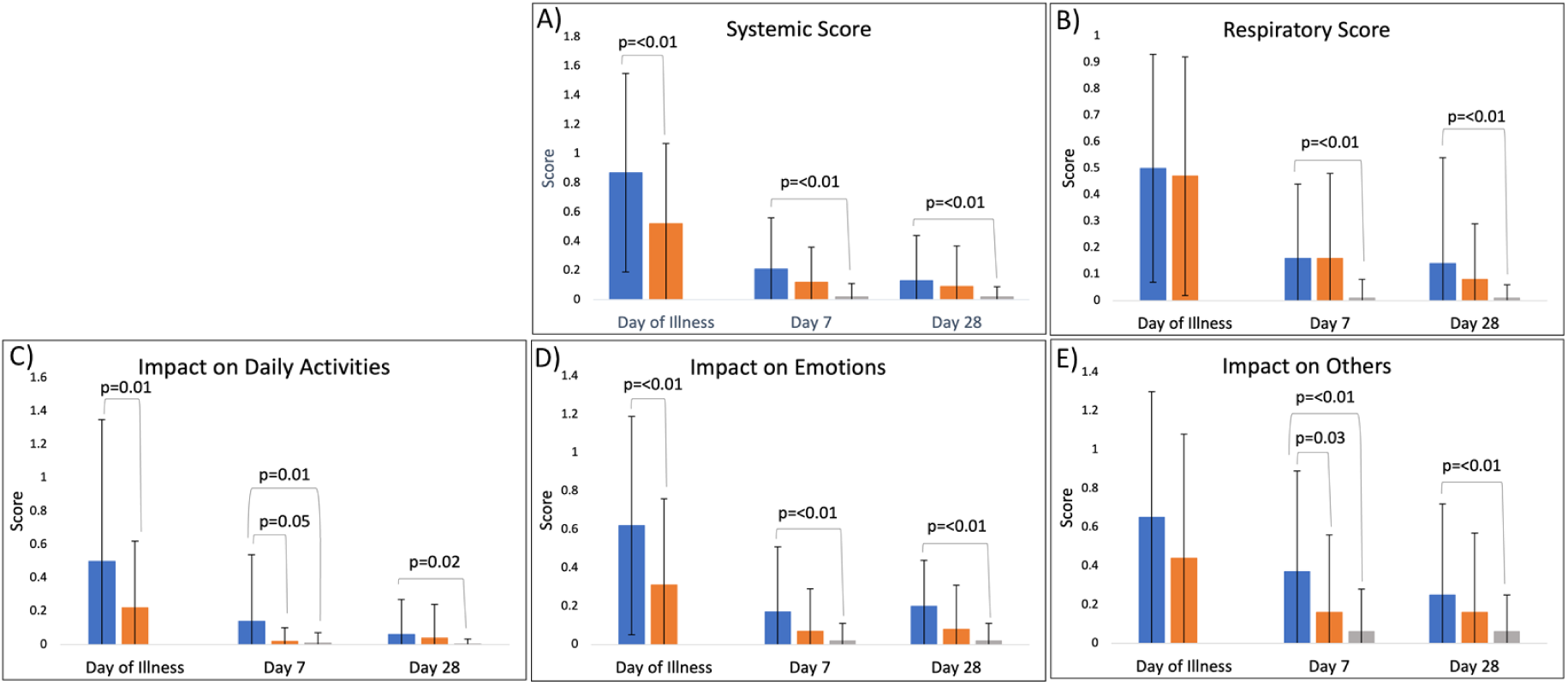
Flu-iiQ severity scores (range: 0-3), by subdomain, for workers with SARS-CoV-2-positive influenza-like illness (ILI), SARS-CoV-2-negative ILI, and asymptomatic controls. Higher score indicates greater clinical severity (Panels A and B) or greater negative impact on well-being (Panels C-E). Significant differences (p<0.05) are identified within each group. Blue=SARS-CoV-2-positive ILI, orange=SARS-CoV-2-negative ILI, gray=asymptomatic controls.

### Economic Outcomes

Compared to SARS-CoV-2-negative workers with ILI, SARS-CoV-2 positive workers had greater self-reported lost income (median US$ 127.1 vs $0, p<0.01), and combined (healthcare, transportation, lost wages) total cost (US$ 147.9 vs $12.7, p<0.01) at day 7 (reported over the preceding 2 weeks) (**Figure 5**); workers with SARS-CoV-2 also had more days of work absence (p<0.01), with most (81.8%) having >5 days of work absence. Household expenditures on fruits/vegetables were higher at day 7 for SARS-CoV-2-positive workers vs test-negatives with ILI (US$ 19.5 vs $13.0, p<0.01); differences in all other household expenditures between SARS-CoV-2 test-positive and test-negative workers were not statistically significant.

**Figure 5.**
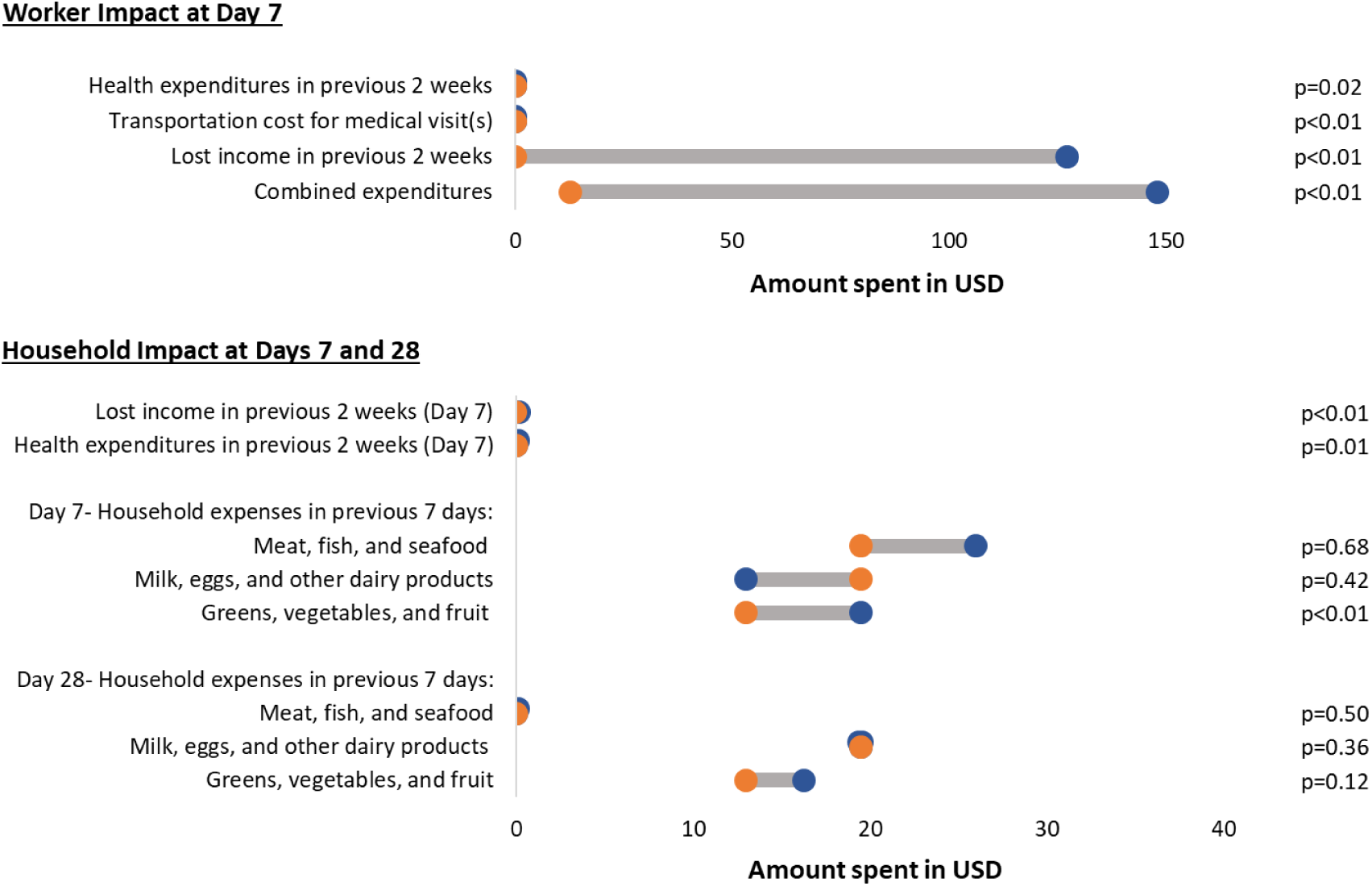
Differences in expenditures between SARS-CoV-2-positive and -negative workers with influenza-like illness (ILI). Workers with SARS-CoV-2-positive ILI (dark blue circle) reported significantly greater lost income and combined expenditures related to their illnesses in the week following their illness than SARS-CoV-2-negative workers with ILI (orange circle).

## DISCUSSION

As of October 10 2021, Guatemalan plantation workers in this prospective cohort study experienced a substantial burden of acute respiratory illness during the COVID-19 pandemic, of which one in four tested positive for SARS-CoV-2; those with COVID-19 had greater disease severity, absenteeism, and economic losses than workers with SARS-CoV-2-negative ILI.

Similar to limited data from the U.S. (1), plantation workers in Guatemala were at risk for SARS-CoV-2 infection (3.1 cases/100 P-Y) throughout the 2020-21 season. Compared to other members of their households, the plantation workers were nearly always the index symptomatic case. These findings, along with the critical role agricultural workers play in Guatemalan and global food security (4, 6), lend support to the prioritization of vaccinating plantation workers against COVID-19.

Although preliminary, our findings suggest COVID-19 illness was associated with greater overall clinical severity and impairment, which persisted at 7- and 28-days post-illness compared to non-SARS-CoV-2 ILI cases. COVID-19 symptoms were consistent with those reported elsewhere, with higher frequencies of anosmia and dysgeusia and prolonged fever differentiating COVID-19 from other ILI cases. Interestingly, COVID-19 was significantly associated with irritability and difficulty concentrating, consistent with post-acute sequelae of SARS-CoV-2 (PASC, or “long COVID”) (32, 33). The irritability and inability to concentrate, which persisted in some workers at both 7 and 28 days, may place workers at risk, for example, when using machetes to harvest bananas and when operating heavy equipment. The Flu-iiQ well-being scores, which include socio-emotional and functional activities, generally indicated more severe illness among workers with COVID-19 compared to workers with other ILI at the time of diagnosis and day 7, with a non-significant trend at 28 days. It is unknown to what extent symptoms or sequelae persist beyond 28 days in this population.

Plantation workers in this cohort experienced a significant economic impact from COVID-19. Self-reported data suggest a significant difference in absenteeism, lost earnings, and total costs between COVID-19 and other ILI cases. Median monthly household income (US$ 363.2), already just below the mean basic monthly food basket price in Guatemala (US$ 386.3), was reduced significantly in workers with COVID-19 (median lost income: US$ 127.1, median total cost of illness: $147.9), placing these households at increased risk for food insecurity and economic hardship. Notably, economic insecurity is one of the primary drivers of emigration from Guatemala (34, 35), and thus the economic impact and policy implications of COVID-19 on these plantation workers and their households, as well as others in similar settings, may extend beyond the Guatemalan border.

While SARS-CoV-2 was the most frequently detected respiratory pathogen among workers with ILI, we detected no cases of influenza and only 6 cases of RSV. Influenza and RSV both circulate year-round in Guatemala and comprise a substantial proportion of ILI cases in population-based studies in Central and South America (36-39). The lower incidence observed in our cohort suggests mitigation strategies–primarily the closing of schools, mask use, and some level of physical distancing–may have been effective in limiting some transmission of influenza and RSV. The observation of rhino/enterovirus and seasonal coronaviruses (NL63, OC43, and N229E) cases in a subset of our cohort is consistent with other reports, (40, 41) though the reasons for these detections despite physical distancing measures merit further study. The AGRI cohort and similar studies will provide important observations on the effectiveness of population-based preventive measures such as vaccines on the burden of respiratory pathogens. Also important, our data demonstrate that syndromic surveillance in the workplace is a feasible population-based approach to rapidly characterize an emerging pathogen.

The AGRI study design has some inherent strengths and limitations. Though the study includes weekly visits to worksites to identify symptomatic ILI cases, it still requires some level of self-reporting to study personnel, and therefore may underestimate incidence. Workers with laboratory-confirmed SARS-CoV-2 are required to isolate and may be incentivized to under-report illness to avoid lost wages, thus providing a bias towards lower incidence and more severe cases of disease being reported. Required isolation likely increases duration of absenteeism in workers who are SARS-CoV-2-positive, though it still reflects the consequences of COVID-19 in this population. Self-reported study outcomes are also subject to recall bias, which we aimed to minimize by including controls with similar follow-up. Laboratory test results are provided to the worker when available, and thus self-reported outcomes may be impacted by diagnostic bias. We did not perform pathogen testing on controls. We relied on an antigen test for detection of SARS-CoV-2 infection and an ELISA assay for anti-nucleocapsid IgG, which may have decreased performance compared to PCR and virus neutralization assays, respectively; future studies will compare these approaches. Future studies will also include company-reported data, which will provide a more objective assessment of wages, allowing us to compare self- and company-reported metrics. Finally, to decrease the risk of healthy worker bias (42), the study collects post-acute (28-day) outcomes on all ILI cases and will ultimately measure loss of employment (using company data) as an outcome measure of ILI.

In conclusion, preliminary data from the AGRI cohort suggest a significant clinical and socioeconomic impact of respiratory illnesses, especially COVID-19, on plantation workers in Guatemala. The study demonstrates the feasibility and value of conducting workforce-based syndromic surveillance during epidemic activity and uses several innovative approaches to measure disease outcomes in the acute and post-acute setting, such as the implementation of active surveillance and molecular diagnostics within a large banana plantation and the inclusion of company-reported economic measures. It also provides a more comprehensive assessment of how communicable diseases economically impact an essential, yet vulnerable, workforce population and their households. Given the high clinical and economic burden of COVID-19 disease among plantation workers in our cohort, and their likely role in household transmission, our results support the prioritization of people working in the agricultural sector for vaccination against COVID-19, potentially through the workplace.

## Data Availability

All data produced in the present work are contained in the manuscript

## FOOTNOTES

## Acknowledgements

We thank the following for their contributions to this research: CU Trifinio Research Team, the CU Center for Global Health administration, Dr. Daniel Jernigan and the Centers for Disease Control and Prevention, AgroAmerica, Margot Charette at TEPHINET for assistance with the mapping, CSU Veterinary Diagnostic Laboratory for use of facilities and the CSU Office of the Vice President for Research for funding instrumentation used in this project. We thank Richard Osborne and Measured Solutions for the use of the Flu-iiQ™ instrument and interpretation. We thank the Trifinio community research subjects.

## CDC Disclaimer

The findings and conclusions in this report are those of the authors and do not necessarily represent the official position of the Centers for Disease Control and Prevention (USA).

## Author Roles

DO, MC, ML, FH, HM, KE, LSN, EJA: conceptualization, methodology, investigation, writing. GG, EB, ANC, NR, KA, GM, GAB, DB, KL, MS, KQ, GE: methodology, investigation, administration. ML, MG, JM, ANC, CI, LMD, EZ, EAB: analysis, visualization, writing. DO, MC, EJA: supervision, administration.

## Conference Presentation

Abstracts was presented at ISIRV 2021 Oct 19-21, 2021 and ASTMH, Nov 17-21, 2021.

## Supplementary Tables

**Supplementary Table 1:** Absenteeism Characteristics, August 31 2020-February 19 2021.

| Characteristics | Workers with<br>no absence<br>(n=695) | Workers with<br>any absence<br>(n=736) | p-value* |
| --- | --- | --- | --- |
| **Worker Demographics** |  |  |  |
| Age: mean (SD) | 32.7 (9.1) | 30.0 (8.0) | <0.01 |
| Male sex, n (%) | 611 (87.9) | 593 (80.6) | <0.01 |
| Ethnicity: Ladino | 312 (44.9) | 315 (42.8) |  |
| Indigenous | 47 (6.8) | 50 (6.8) | 0.82 |
| other | 1 (0.1) | 2 (0.3) |  |
| don't know | 335 (48.2) | 369 (50.1) |  |
| **Worker Health, n (%)** |  |  |  |
| Asthma | 1 (0.1) | 7 (1.0) | 0.04 |
| Pulmonary disease | 4 (0.6) | 5 (0.7) | 0.80 |
| Kidney disease | 27 (3.9) | 22 (3.0) | 0.35 |
| Cardiovascular disease | 10 (1.4) | 10 (1.4) | 0.90 |
| Diabetes | 13 (1.9) | 7 (1.0) | 0.14 |
| Blood disorder (sickle cell) | 8 (1.2) | 14 (1.9) | 0.25 |
| Neurologic disease (stroke) | 3 (0.4) | 5 (0.7) | 0.53 |
| Liver disease | 5 (0.7) | 10 (1.4) | 0.24 |
| Obesity (BMI≥30 kg/m<sup>2</sup>), n=773 | 67 (15.6) | 15 (4.4) | <0.01 |
| Taking medications? | 83 (11.9) | 95 (12.9) | 0.57 |
| Ever received influenza vaccine | 50 (7.2) | 41 (5.6) | 0.21 |
| **Work Conditions** |  |  |  |
| # of absences/person, median (IQR) | 0 (0, 0) | 5.3 (2, 7) | <0.01 |
| Type of Work, n (%) |  |  |  |
| Administration | 43 (6.2) | 0 (0) |  |
| Field worker | 42 (66.5) | 528 (71.7) | <0.01 |
| Field manager | 54 (7.8) | 3 (0.4) |  |
| Packer (factory worker) | 120 (17.3) | 205 (27.9) |  |
| Factory manager | 16 (2.3) | 0 (0) |  |
| How long worked at farm, n (%) |  |  |  |
| ≤2 years | 346 (49.8) | 453 (61.5) | <0.01 |
| 3-4 years | 104 (14.9) | 113 (15.4) |  |
| ≥5 years | 245 (35.3) | 170 (23.1) | |
| Monthly income, \$USD, median (IQR) | 337.2 (311.3, 389.1) | 337.2 (311.3, 363.2) | <0.01 |
| **Household Conditions** |  |  |  |
| # adults in house, mean (SD) | 3.2 (1.6) | 3.4 (1.8) | 0.01 |
| # children in house, mean (SD) | 2.3 (1.6) | 2.5 (1.7) | <0.01 |
| Concern about food insecurity in last year, n (%) | 401 (57.7) | 464 (63.0) | 0.04 |
| Lacked money for food in the last 12 months, n (%) | 506 (72.8) | 505 (68.6) | 0.08 |
| Household monthly income, \$USD, median (IQR) | 363.2 (324.3, 466.9) | 376.1 (324.3, 466.9) | 0.06 |
| \$USD spent in the last 7 days, median (IQR): | | | |
| Meat, fish, and seafood | 22.7 (13.0, 32.4) | 19.5 (13.0, 38.9) | 0.68 |
| Milk, eggs, and other dairy products | 13.0 (9.1, 24.9) | 15.6 (9.7, 25.9) | 0.07 |
| Greens, vegetables, and fruit | 13.0 (6.5, 19.5) | 13.0 (6.5, 19.5) | 0.83 |
| Alcoholic drinks and tobacco | 0 (0, 0) | 0 (0, 0) | 0.16 |

\*p-values are from t-tests for continuous variables, and chi-square test for categorical variables.
Abbreviations: SD=standard deviation, BMI=body mass index, IQR=interquartile range, USD=US dollars

**Supplementary Table 2.** Comparing Self-Reported Outcomes in ILI vs Asymptomatic Controls.

|  |  |  |  |  |  |  |  |
| --- | --- | --- | --- | --- | --- | --- | --- |
|  | **Day 0 ILI**<br>(n=169) | **D7 ILI**<br>(n=157) | **D7 Control**<br>(n=623) | **p-value\*** | **D28 ILI**<br>(n=149) | **D28 Control**<br>(n=588) | **p-value\*** |
| ***Clinical Symptom*** |  |  |  |  |  |  |  |
| # days of cough (if cough), mean (SD) | 3.5 (2.1) | 5.3 (2.6) | 3.4 (2.6) | 0.05 | 5.0 (3.5) | 3.7 (2.0) | 0.32 |
| # days of fever (if fever), mean (SD) | 2.6 (1.8) | 3.6 (2.6) | 1.5 (0.6) | <0.01 | 2.6 (2.3) | 3.5 (3.0) | 0.55 |
| Have you felt the following (last 24 hours), n (%): |  |  |  |  |  |  |  |
| Fever | 96 (57.5) | 14 (9.0) | 4 (0.7) | <0.01 | 8 (5.4) | 3 (0.5) | <0.01 |
| Nasal Congestion | 60 (35.9) | 23 (14.7) | 16 (2.7) | <0.01 | 9 (6.1) | 6 (1.1) | <0.01 |
| Myalgia | 73 (43.7) | 14 (9.0) | 23 (3.9) | 0.01 | 11 (7.5) | 11 (2.0) | <0.01 |
| Headache | 89 (53.3) | 37 (23.7) | 22 (3.7) | <0.01 | 25 (17.0) | 14 (2.5) | <0.01 |
| Cough | 69 (41.3) | 22 (14.2) | 2 (0.34) | <0.01 | 10 (6.8) | 1 (0.2) | <0.01 |
| Sore Throat | 66 (39.5) | 20 (12.8) | 7 (1.2) | <0.01 | 12 (8.2) | 10 (1.8) | <0.01 |
| Dysgeusia | 51 (30.5) | 15 (9.6) | 2 (0.3) | <0.01 | 6 (4.1) | 2 (0.4) | <0.01 |
| Expectoration | 62 (37.1) | 30 (19.2) | 4 (0.7) | <0.01 | 12 (8.2) | 2 (0.4) | <0.01 |
| Fatigue | 58 (34.7) | 19 (12.2) | 18 (3.0) | <0.01 | 11 (7.5) | 19 (3.4) | 0.03 |
| Anosmia | 40 (24.0) | 13 (8.3) | 1 (0.2) | <0.01 | 6 (4.1) | 2 (0.4) | <0.01 |
| Loss of Appetite | 49 (29.3) | 10 (6.4) | 6 (1.0) | <0.01 | 3 (2.0) | 7 (1.3) | 0.44 |
| Dyspnea | 41 (24.7) | 10 (6.4) | 1 (0.2) | <0.01 | 5 (3.4) | 1 (0.2) | <0.01 |
| Neck Pain | 34 (20.5) | 11 (7.1) | 8 (1.3) | <0.01 | 8 (5.4) | 8 (1.4) | <0.01 |
| Interrupted Sleep | 33 (19.8) | 15 (9.6) | 7 (1.2) | <0.01 | 7 (4.8) | 6 (1.1) | <0.01 |
| Wheezing | 19 (11.5) | 6 (3.9) | 1 (0.2) | <0.01 | 5 (3.4) | 0 (0) | <0.01 |
| ***Well-Being\*\**** |  |  |  |  |  |  |  |
| Have you had difficulty with (last 24 hrs), n (%): |  |  |  |  |  |  |  |
| Getting out of bed | 44 (26.4) | 10 (6.4) | 7 (1.2) | <0.01 | 7 (4.8) | 6 (1.1) | <0.01 |
| Preparing meals... | 18 (10.8) | 4 (2.6) | 3 (0.5) | 0.04 | 4 (2.7) | 1 (0.2) | 0.01 |
| Performing usual... | 45 (27.0) | 9 (5.8) | 4 (0.7) | <0.01 | 6 (4.1) | 2 (0.4) | <0.01 |
| Leaving the home... | 27 (16.2) | 6 (3.9) | 2 (0.3) | <0.01 | 4 (2.7) | 1 (0.2) | 0.01 |
| Concentrating on... | 43 (25.8) | 9 (5.8) | 7 (1.2) | <0.01 | 7 (4.8) | 1 (0.2) | <0.01 |
| Taking care of... | 31 (18.6) | 4 (2.6) | 3 (0.5) | 0.04 | 6 (4.1) | 1 (0.2) | <0.01 |
| Leave the room | 23 (13.8) | 4 (2.6) | 2 (0.3) | 0.02 | 4 (2.7) | 0 (0) | <0.01 |
| Have you felt the following (last 24 hours), n (%): |  |  |  |  |  |  |  |
| Irritable | 66 (39.5) | 13 (8.3) | 10 (1.7) | <0.01 | 10 (6.8) | 5 (0.9) | <0.01 |
| Defenseless | 40 (24.0) | 12 (7.7) | 9 (1.5) | <0.01 | 6 (4.1) | 4 (0.7) | 0.01 |
| Worried | 59 (35.3) | 23 (14.7) | 13 (2.2) | <0.01 | 15 (10.2) | 20 (3.6) | <0.01 |
| Frustrated | 34 (20.4) | 10 (6.4) | 7 (1.2) | <0.01 | 7 (4.8) | 8 (1.4) | 0.01 |
| People worrying... | 93 (55.7) | 40 (25.6) | 56 (9.4) | <0.01 | 29 (19.7) | 69 (12.4) | 0.02 |
| Being a burden | 47 (28.1) | 20 (12.8) | 12 (2.0) | <0.01 | 14 (9.5) | 15 (2.7) | <0.01 |
| People being... | 38 (22.8) | 16 (10.3) | 16 (2.7) | <0.01 | 15 (10.2) | 15 (2.7) | <0.01 |
| Needing to depend... | 42 (25.2) | 19 (12.2) | 20 (3.4) | <0.01 | 16 (10.9) | 19 (3.4) | <0.01 |
| People having to do extra... | 37 (22.2) | 21 (13.5) | 22 (3.7) | <0.01 | 16 (10.9) | 20 (3.6) | <0.01 |
| ***Flu-iiQ Severity Scores*** |  |  |  |  |  |  |  |
| Systemic Score | 0.60 (0.47) | 0.14 (0.28) | 0.02 (0.09) | <0.01 | 0.10 (0.28) | 0.02 (0.07) | <0.01 |
| Respiratory Score | 0.46 (0.44) | 0.15 (0.11) | 0.01 (0.07) | <0.01 | 0.09 (0.27) | 0.01 (0.05) | <0.01 |
| Impact on daily activities | 0.28 (0.49) | 0.05 (0.22) | 0.01 (0.06) | 0.01 | 0.04 (0.19) | 0.004 (0.03) | 0.02 |
| Impact on Emotions Score | 0.39 (0.51) | 0.11 (0.28) | 0.02 (0.09) | <0.01 | 0.07 (0.22) | 0.02 (0.09) | <0.01 |
| Impact on Others Score | 0.49 (0.65) | 0.21 (0.43) | 0.06 (0.22) | <0.01 | 0.17 (0.41) | 0.06 (0.19) | <0.01 |
| **Epidemiology** |  |  |  |  |  |  |  |
| # in house w/ similar illness (last 2 weeks) | 0.17 (0.45) | 0.21 (0.52) | 0.01 (0.10) | <0.01 | 0.12 (0.38) | 0.01 (0.15) | <0.01 |
| Index case in house, n (%) |  |  |  |  |  |  |  |
| Self | 145 (86.8) | 142 (91.0) | 1 (14.3) |  | 135 (91.8) | 3 (42.9) |  |
| Spouse | 5 (3.0) | 2 (1.3) | 2 (28.6) |  | 2 (1.4) | 1 (14.3) |  |
| Parent | 2 (1.2) | 2 (1.3) | 1 (14.3) | <0.01 | 1 (0.7) | 0 (0) | <0.01 |
| Sibling | 6 (3.6) | 5 (3.2) | 0 (0) |  | 6 (4.1) | 0 (0) |  |
| Cousin | 1 (0.6) | 1 (0.6) | 0 (0) |  | 0 (0) | 0 (0) |  |
| Child | 4 (2.4) | 1 (0.6) | 3 (42.9) |  | 2 (1.4) | 2 (28.6) |  |
| *p-values are from t-tests for continuous variables, and chi-square test for categorical variables. |  |  |  |  |  |  |  |
| ** Items truncated per Flu-iiQ licensing agreement; full items available from the author. |  |  |  |  |  |  |  |
| Abbreviations: ILI = influenza-like illness case, D7 = day 7, D28 = day 28. |  |  |  |  |  |  |  |

